# Does a probiotic (L. reuteri) lozenge taken twice daily over 3-4 weeks reduce probing pocket depth in patients with chronic periodontitis after 3 months? A systematic review of clinical trials (Protocol)

**DOI:** 10.1101/2023.04.24.23289012

**Authors:** Steffen Mickenautsch, Stefan Rupf, Veerasamy Yengopal

**Affiliations:** Review Centre for Health Science Research, 84 Concorde Road East, Bedfordview/Johannesburg, 2008, South Africa; Department of Community Dentistry, School of Oral Health Sciences, Faculty of Health Sciences, University of the Witwatersrand, 7 York Rd, Parktown/Johannesburg, 2193, South Africa; Synoptic Dentistry, Saarland University, Building 73, 66421, Homburg, Germany; Faculty of Dentistry, University of the Western Cape, Francie van Zijl Avenue, Tygerberg/Cape Town, 7505, South Africa

## Abstract

**Introduction:** Chronic periodontitis is a slow progressing, multifactorial inflammatory disease of the periodontium that may lead to its destruction, which is detectable as increasing probing pocket depth (PPD), subsequent tooth mobility and tooth loss. The purpose of this systematic review is to update and appraise the current trial evidence to the question do probiotic (L. reuteri) lozenge taken twice daily over 3-4 weeks reduce probing pocket depth in patients with chronic periodontitis after 3 months.

**Methods and analysis:** We will conduct reference checks of previous systematic review and trial reports to the topic. PubMed, Scopus, Cochrane library and the Directory of Open Access Journals (DOAJ) will be searched. All selected trial reports will be independently appraised by two reviewers, using the CQS-2B trial appraisal tool. Meta-analysis will be conducted using a random effect model with inverse variance method, stratified according to CQS-2B corroboration levels (C1 – 4). The I^2^ – test with 95% Confidence Interval will be used to establish whether any statistical heterogeneity between datasets exist. Sensitivity analysis will be conducted for meta-analysis results of trials that were rated with 1-score at all four CQS-2B appraisal criteria, by excluding trials in which: Patients were smokers; Patients were Type II diabetics; Adjunctive antibiotic therapy was provided. For meta-analyses including data of at least four trials, which have been rated with a 1-score for all four appraisal criteria, the results will be further statistically tested for possible selection bias. Publication bias risk will be quantitatively assessed by use of Egger’s regression.

**Ethics and dissemination:** Ethical approval is not required for literature-based studies. The results will be disseminated as a prior preprint version and subsequent peer-reviewed publication.

## Introduction

Chronic periodontitis is a slow progressing, multi-factorial inflammatory disease of the periodontium that may lead to its destruction, which is detectable as increasing probing pocket depth (PPD), subsequent tooth mobility and tooth loss [1]. Treatment in its early stage comprises of non-surgical periodontal therapy (NSPT), particularly of scaling and root planning (SRP) [2].

A comprehensive systematic review by Ausenda et al (2023) appraised the clinical evidence regarding the possible beneficial effect of the adjunctive use of a probiotic for chronic periodontitis treatment. The authors appraised the clinical evidence for the type of probiotic application (via lozenge, capsule or other); application frequency (1- or 2-times per day); type of probiotic (Lactobacillus reuteri, others), type of measured outcome (probing pocket depth – PPD reduction, clinical attachment level – CAL gain) and the length of follow-up period (< 3, 3 – 12, > 12 months) [2]. The result of the systematic review showed that, beside the application of NSPT and, the intake of a probiotic lozenge, containing L. reuteri, twice daily for a period of 3-4 weeks in comparison to placebo was associated with the highest treatment benefit in terms of a statistically significant PPD reduction after a minimum period between 3 - 12 months. The point estimates of the mean differences (MD) with 95% Confidence interval (CI) for PPD reduction was higher and thus even more promising than that of the CAL gain for the same type of probiotic, application type and frequency [2]. An earlier second systematic review by Song and Liu (2020) reported similar results [3].

Both systematic reviews appraised clinical trials using the first version of Cochrane’s Risk of Bias (RoB) tool [4]. While most of the trials were judged of low-bias risk, some were rated of moderate and high risk, due to lack of adequate randomisation and blinding [2,3]. However, both systematic reviews based their review conclusions solely on the established trial data, without neither stratification by overall risk-of-bias judgment nor any other form of quantitatively integrating the established bias risk into their clinically relevant conclusions. In addition, the first RoB version has been found to have low inter-rater reliability: kappa 0.54 (95% confidence interval (CI): 0.29–0.79) [5], kappa 0.50 (95% CI: 0.36–0.63); and low agreement across reviewer pairs: kappa 0.37 (95% CI: 0.19–0.55) [6], which carries the high risk that the results, established in both systematic reviews, may be subjective and not reviewer independent. For that reason, potential high bias risk may have affected an overestimation of the systematic review results and subsequently its clinically relevant conclusions.

In contrast, it has been shown that the Composite Quality Score (CQS) for the appraisal of prospective, controlled clinical therapy trials has a high interrater reliability: Brennan-Prediger coefficient (BPC) of 0.95; 95% CI: (0.87–1.00), which compared favourably to that of the first RoB tool version, with most of the differences between the RoB and the CQS being statistically significant (p < 0.05) in favour of the CQS [7]. In addition, the latest CQS version (CQS-2B), besides its high inter-rater reliability [8] has been established on a rigorous evidence base [9, 10], sound epistemic principles [11] and is recommended to be applied together with stratification by overall risk-of-bias judgment of the established effect estimates [12]

The purpose of this systematic review is to update and appraise the current trial evidence to the question: Does a probiotic (L. reuteri) lozenge taken twice daily over 3-4 weeks reduce probing pocket depth in patients with chronic periodontitis after 3 months?

## Materials and methods

The protocol of this systematic review will be made available online and registered with the National Institute for Health Research PROSPERO, International Register of Systematic Reviews.

### Participants, intervention, comparison, outcome and study design (PICOS)

#### Participants (P)

Adult patients suffering from chronic periodontitis.

#### Intervention (I)

NSPT with additional administration of a probiotic lozenge containing L. reuteri, taken two times per day for a 3 – 4 week (21 – 28 day) period.

#### Comparison (C)

NSPT with placebo, taken two times per day for a 3 – 4 week (21 – 28 day) period.

#### Outcome (O)

Changes in pocket probing (PPD) depth after a minimum time period of three months (up to a maximum of 12 months) after start of treatment. The PPD is defined as the difference between the gingival margin and the bottom of the periodontal pocket, measured with a periodontal probe in millimetres (mm). The measure of effect for the PPD changes will be the mean difference (MD) with 95% Confidence interval (CI).

#### Study design (S)

Prospective, controlled clinical therapy trials with parallel group design.

### Systematic literature search

References checks of the two previous systematic review reports [2, 3], as well as of identified trials, for suitable trial reports will be conducted. The search period for the systematic review by Song and Liu (2020) was between 2009 – 2019 [3] and the search cut-off date for the systematic review by Ausenda et al (2023) was March 05, 2020) [2]. Therefore, it is assumed that the systematic literature search by the two reviews have identified all relevant trial reports published prior March 05, 2020.

In addition to the reference check, the following databases: PubMed, Scopus, Cochrane library and the Directory of Open Access Journals (DOAJ) will be searched using the string of search terms: chronic periodontitis AND lactobacillus reuteri. The search in PubMed will be limited between ‘March 05, 2020 and present’, the search in Scopus between ‘2020 – present’ and the search in the Cochrane library between ‘March 2020 – present’.

One reviewer will conduct the searches by screening citation titles and abstracts and retrieve the full-text articles. A second reviewer will independently verify the retrieved trials reports for eligibility. Any disagreements will be resolved via discussion and consensus.

### Trial selection criteria

Published trial reports in any publication languages that comply with all the following criteria will be selected:

i. Prospective, controlled clinical therapy trial;
ii. Trial characteristics in line with specified PICOS;
iii. Trial report published in full;
iv. Computable continuous data for test and control group reported, including: total number of subjects, mean PPD values with standard deviation (SD) or standard error (SE).

Trial reports that during the review process are found not to comply with all criteria will be excluded. Where more than one report exists per trial the one with the most recent publication date will be selected.

### Data extraction from accepted trials

All trial reports that are deemed relevant during the systematic literature search will be traced in full copy and the following information extracted:

i. Full reference details;
ii. Basic trial characteristics:
  a. Number of patients enrolled at baseline per intervention group;
  b. Mean patient age with SD per intervention group;
  c. Patient gender distribution (Male/Female) in %;
  d. Patients are smokers (Yes/No);
  e. Patients are Type II diabetics (Yes/No);
  f. Adjunctive antibiotic therapy provided (Yes/No);
iii. Computable data per intervention group.

One reviewer will extract all information and enter them into an MS Excel file. A second reviewer will double-check the extracted data and corrected possible entry errors. Any disagreements will be resolved via discussion and consensus.

### Main data analysis

Any extracted standard errors (SE) will be converted into standard deviations (SD). Meta-analysis will be conducted using a random effect model with inverse variance method, stratified according to CQS-2B corroboration levels (C1 – 4).

The I^2^ –test with 95% CI will be used to establish whether any statistical heterogeneity between datasets exist. Thresholds for I^2^ point estimates (in %) and its upper confidence values will be used in order to interpret the test results: 0–40% = might not be important; 30–60% = may represent moderate heterogeneity; 50–90% = may represent substantial heterogeneity; 75–100% = considerable heterogeneity [13].

### Sensitivity analysis

Sensitivity analysis will be conducted for meta-analysis results of trials that were rated with 1-score at all four CQS-2B appraisal criteria, by excluding trials in which:

a. Patients were smokers;
b. Patients were Type II diabetics;
c. Adjunctive antibiotic therapy was provided.

The results of the sensitivity analysis will be compared to that of the main analysis, in order to ascertain, whether smoking, Type II diabetes and antibiotic therapy had any possible confounding effect on the established main results.

### Basic assessment of bias risk

All selected trial reports will be independently appraised by two reviewers, using the CQS-2B tool (Table 1). Any disagreements between reviewers will be resolved by discussion and consensus.

**Table 1.** CQS-2B appraisal criteria.

|  |  |  |  |
| --- | --- | --- | --- |
| Criterion I | 'Randomisation' for allocation to treatment groups is in some form reported in the text | Yes | No |
| Criterion II | Any assurance that the patient allocation to treatment groups according to the random sequence was applied by an independent agent or agency, not otherwise involved in the trial, is in some form reported in the text |  |  |
| Criterion III | Double-blinding or the blinding of at least two out of the three groups: trial participants trial personnel and trial outcome assessors in some form reported in the text |  |  |
| Criterion IV | The sample size of any particular treatment group reported in the trial is not less than N = 100 |  |  |

Application of the CQS-2B comprises: (i) binary trial report rating per appraisal criterion (Scores: 0 = No/invalid/falsified, 1 = Yes/corroborated); (ii) multiplication of all criterion scores to an overall appraisal score, and (iii) identification of invalid/falsified trial reports based on a zero overall appraisal score.

During CQS-2B application, several corroboration (C-) levels are recognised. C-levels indicate the number of consecutive criteria that a trial has complied with (e.g. level C2 indicates compliance with Criterion I and II; level C3 indicates compliance with Criterion I, II and III, etc.) [11]. A corroboration level for a particular trial is reached before one criterion is rated with a 0-score or when all criteria are rated with a 1-score; for example, Corroboration level C2: Criterion I and II = 1-score, Criterion III = 0-score; Corroboration level C4: All criteria = 1-score. After a criterion has been rated with a 0-score, the C-level of a trial remains the same even if a following criterion is rated with a 1-score, for example, Corroboration level C2: Criterion I and II = 1-score, Criterion III = 0-score, Criterion IV = 1-score.

An overall 1-score appraisal result will indicate that a trial is ‘corroborated’, which means that during the appraisal process no evidence in support for the assumption that its reported results are compromised by high bias risk has been established. This does not mean that such evidence may not be identified during future appraisals with additional appraisal criteria. For that reason, a corroborated trial will not be assumed to be of ‘low bias risk’ status.

For all allocated 1-scores the appropriate verbatim quotes will be extracted from the trial report and entered into a verbatim table, including page number/column/paragraph number/line number of the trial report.

### Further assessment of bias risk

For meta-analyses including data of at least four trials at CQS-2B corroboration level 4 (i.e. trials which have been rated with a 1-score for all four appraisal criteria), the results will be further statistically assessed for possible selection bias, caused by inadequate allocation concealment, using the statistical test presented by Hicks et al. [14]. In addition, all trials judged at corroboration level 4 will be appraised for any other type of possible error related to their individual trial characteristics.

Publication bias risk will be quantitatively assessed (including trials from all corroboration levels: 1-4) by use of Egger’s regression. Publication bias will be assumed to be present at significance level p < 0.10 [15]. Publication bias will not be assessed if the number of selected trials is < 10.

### Reporting

Reporting of the systematic review will follow PRISMA guidelines. The final report will be made available online as preprint and submitted to a suitable peer-reviewed journal for possible publication.

## Data Availability

All data produced in the present work are contained in the manuscript

## Data availability

All data will be made fully available without restriction as part of the preprint and journal publication.

## References

[1] Papapanou PN, Sanz M, Buduneli N, Dietrich T, Feres M, Fine DH, et al. Periodontitis: consensus report of workgroup 2 of the 2017 world workshop on the classification of periodontal and peri-implant diseases and conditions. J Periodo 2018;89:S173–82.

[2] Ausenda F, Barbera E, Cotti E, Romeo E, Natto ZS, Valente NA. Clinical, microbiological and immunological short, medium and long-term effects of different strains of probiotics as an adjunct to non-surgical periodontal therapy in patients with periodontitis. Systematic review with meta-analysis. Jpn Dent Sci Rev. 2023 Dec;59:62–103.

[3] Song D, Liu XR. Role of probiotics containing Lactobacillus reuteri in adjunct to scaling and root planing for management of patients with chronic periodontitis: a meta-analysis. Eur Rev Med Pharmacol Sci. 2020 Apr;24(8):4495–4505.

[4] Higgins JP, Altman DG, Gotzsche PC, Juni P, Moher D, Oxman AD, Savovic J, Schulz KF, Weeks L, Sterne JA; Cochrane Bias Methods Group; Cochrane Statistical Methods Group (2011) The Cochrane Collaboration’s tool for assessing risk of bias in randomised trials. BMJ 343:d5928.

[5] Hartling L, Bond K, Vandermeer B, Seida J, Dryden DM, Rowe BH (2011) Applying the risk of bias tool in a systematic review of combination long-acting beta-agonists and inhaled corticosteroids for persistent asthma. PLoS ONE 6:e17242.

[6] Hartling L, Hamm MP, Milne A et al (2013) Testing the risk of bias tool showed low reliability between individual reviewers and across consensus assessments of reviewer pairs. J Clin Epidemiol 66:973–981.

[7] Mickenautsch S, Miletić I, Rupf S, Renteria J, Göstemeyer G. The Composite Quality Score (CQS) as a trial appraisal tool: inter-rater reliability and rating time. Clin Oral Investig. 2021; 25:6015–23.

[8] Mickenautsch S, Rupf S, Miletić I, Strähle UT, Sturm R, Kimmie-Dhansay F, Vidosusić K, Yengopal V. Inter-rater reliability of the extended Composite Quality Score (CQS-2) – a pilot study, 23 November 2022, PREPRINT (Version 1) available at Research Square [https://doi.org/10.21203/rs.3.rs-2297364/v1]

[9] Mickenautsch S, Rupf S, Miletić I, Yengopal V. Extension of the Composite Quality Score (CQS) as an appraisal tool for prospective, controlled clinical therapy trials-A systematic review of metaepidemiological evidence. PLoS One. 2022; 17:e0279645.

[10] Mickenautsch S, Yengopal V. Allocation concealment appraisal of clinical therapy trials using the extended Composite Quality Score (CQS-2) – An empirically based update (Preprint), 14 February 2023, PREPRINT (Version 1) available at Research Square [https://doi.org/10.21203/rs.3.rs-2582208/v1].

[11] Mickenautsch S, Rupf S, Miletić I, Yengopal V. The Composite Quality Score (CQS) as an appraisal tool for prospective, controlled clinical therapy trials: rationale and current evidence. Rev Recent Clin Trials. 2023; 18:28–33.

[12] Mickenautsch S, Rupf S, Yengopal V. Application of the Composite Quality Score (CQS-2B) in systematic reviews of prospective, controlled, clinical therapy trials – an exploratory study (Preprint), 23 March 2023, PREPRINT (Version 1) available at Research Square [https://doi.org/10.21203/rs.3.rs-2718841/v1].

[13] Deeks JJ, Higgins JPT, Altman DG (editors). Chapter 10: Analysing data and undertaking meta-analyses. In: Higgins JPT, Thomas J, Chandler J, Cumpston M, Li T, Page MJ, Welch VA (editors). Cochrane Handbook for Systematic Reviews of Interventions version 6.3 (updated February 2022). Cochrane, 2022. Available from https://www.training.cochrane.org/handbook. (Accessed: April 14, 2023).

[14] Hicks A, Fairhurst C, Torgerson DJ. A simple technique investigating baseline heterogeneity helped to eliminate potential bias in meta-analyses. J Clin Epidemiol. 2018; 95:55–62.

[15] Egger M, Davey Smith G, Schneider M, Minder C (1997) Bias in meta-analysis detected by a simple, graphical test. BMJ 315:629–634.

